# Parkinson’s Disease motor and non-motor progression models emerge from pathway-level transcriptomics

**DOI:** 10.64898/2026.02.25.26346261

**Authors:** Jaime Ñíguez, Antonio Guillén, Guillermo Rocamora, Huw Morris, Mina Ryten, José T. Palma, Juan A. Botia, Ana-Luisa Gil-Martínez

## Abstract

**Background:** Prognosis and therapeutic management in Parkinson’s disease remain challenging due to the disease’s heterogeneous progression and symptom presentation and lack of reliable biomarkers to predict individual disease trajectories.

**Objective:** To determine whether baseline blood transcriptomes, analyzed through biologically defined pathway gene sets, contain signatures that distinguish distinct motor and non-motor progression trajectories in Parkinson’s disease.

**Methods:** Using data from the Parkinson’s Progression Markers Initiative cohort, we developed a pathway-based computational framework to derive individualized molecular severity scores from baseline blood transcriptomic profiles by integrating pathway-level gene expression with longitudinal clinical data. Severity indices for motor and non-motor features established domain-specific progression trajectories of sporadic Parkinson’s disease. Machine learning models were trained to predict patient trajectory membership from baseline transcriptomics. Findings were validated in genetic subcohorts and externally in the Parkinson’s Disease Biomarkers Program cohort.

**Results:** Molecular severity scores were associated with key clinical features. Analysis of score changes revealed two non-motor and two motor progression groups, each characterized by specific gene signatures (20 genes for non-motor; 121 for motor). From baseline transcriptomic data, we accurately predicted an individual’s trajectory group (0.87 for motor progression). The framework demonstrated high generalizability across independent and genetic cohorts, producing clinically coherent profiles.

**Conclusions:** Our analysis reveals that baseline blood transcriptomic profiles delineate motor and non-motor progression trajectories in sporadic Parkinson’s disease. The results are consistent with prior findings and may contribute to the identification of novel biomarkers, thereby informing and potentially optimizing the design of clinical trials aimed at modifying disease progression.

## Introduction

Parkinson’s disease (PD) is a progressive neurodegenerative disorder characterized by substantial heterogeneity in both symptom burden and disease course. While bradykinesia, rigidity, and tremor constitute core motor features, patients show wide variability in the severity and rate of progression of motor and non-motor manifestations, including cognitive impairment and autonomic dysfunction.^1,2^ Understanding this heterogeneity requires moving beyond static, cross-sectional subtyping to identify the biological mechanisms that underlie differential patterns of longitudinal progression.^3^

Large, deeply phenotyped longitudinal cohorts, including the Parkinson’s Progression Markers Initiative^4^ (PPMI) and the Parkinson’s Disease Biomarkers Program^5^ (PDBP), now harmonized within the Accelerating Medicines Partnership–Parkinson’s Disease (AMP-PD) framework, provide an unprecedented opportunity to investigate these dynamics.

Blood-based transcriptomic profiling offers a minimally invasive and scalable window into systemic molecular alterations in PD. Prior studies have consistently implicated pathways related to immune dysregulation and mitochondrial dysfunction, and oxidative stress. However, these studies have largely focused on discriminating patients from healthy controls, offering limited insight into patient-specific heterogeneity or longitudinal progression dynamics.^6–9^ A critical knowledge gap persists in linking temporal variation within these biologically defined pathways to individual differences in symptom severity and progression rates over time.^10^

In this study, we develop a computational framework to link longitudinal blood transcriptomic profiles with dynamic motor and non-motor progression in PD. Specifically, we (i) stratify patients based on pathway-level gene expression patterns, (ii) integrate multiple clinical covariates to derive individualized transcriptomic severity scores, (iii) model progression using longitudinal changes in these severity estimates, and (iv) identify molecular pathways that distinguish distinct progression trajectories. By integrating pathway-informed transcriptomics with longitudinal clinical data, our approach aims to provide insight into the biological heterogeneity underlying differential disease progression in Parkinson’s disease.

## Materials and Methods

### Study design and cohorts

We conducted a longitudinal transcriptomic and clinical analysis using data from the Parkinson’s Progression Markers Initiative (PPMI) cohort^4^. Primary analyses included sporadic Parkinson’s disease (sPD) patients with available whole-blood RNA sequencing and complete clinical assessments at baseline (M0) and at 12-, 24-, and 36-month follow-up visits (M12, M24, M36).

Framework robustness and generalizability were assessed through internal replication in PPMI genetic subcohorts (*SNCA*, *GBA*, and *LRRK2* mutation carriers) as well as in healthy controls (HC). Independent external replication was conducted using data from the PDBP cohort^5^, which provided transcriptomic and clinical data at baseline and at 6-, 12-, 18-, and 24-month visits (M6, M12, M18, M24). Identical preprocessing and analytical steps were applied to both cohorts, with adaptations limited to cohort-specific visit schedules.

### Transcriptomic data preprocessing

Transcriptomic data from the PPMI and PDBP cohorts were processed using a standardized pipeline. Genes with near-zero variance or low expression were excluded using an elbow-based threshold applied to the gene retention curve (Supp. Fig 1.), ensuring stable expression in at least 80% of samples. This procedure yielded 15584 genes in PPMI and 17494 genes in PDBP.

**Figure 1.**
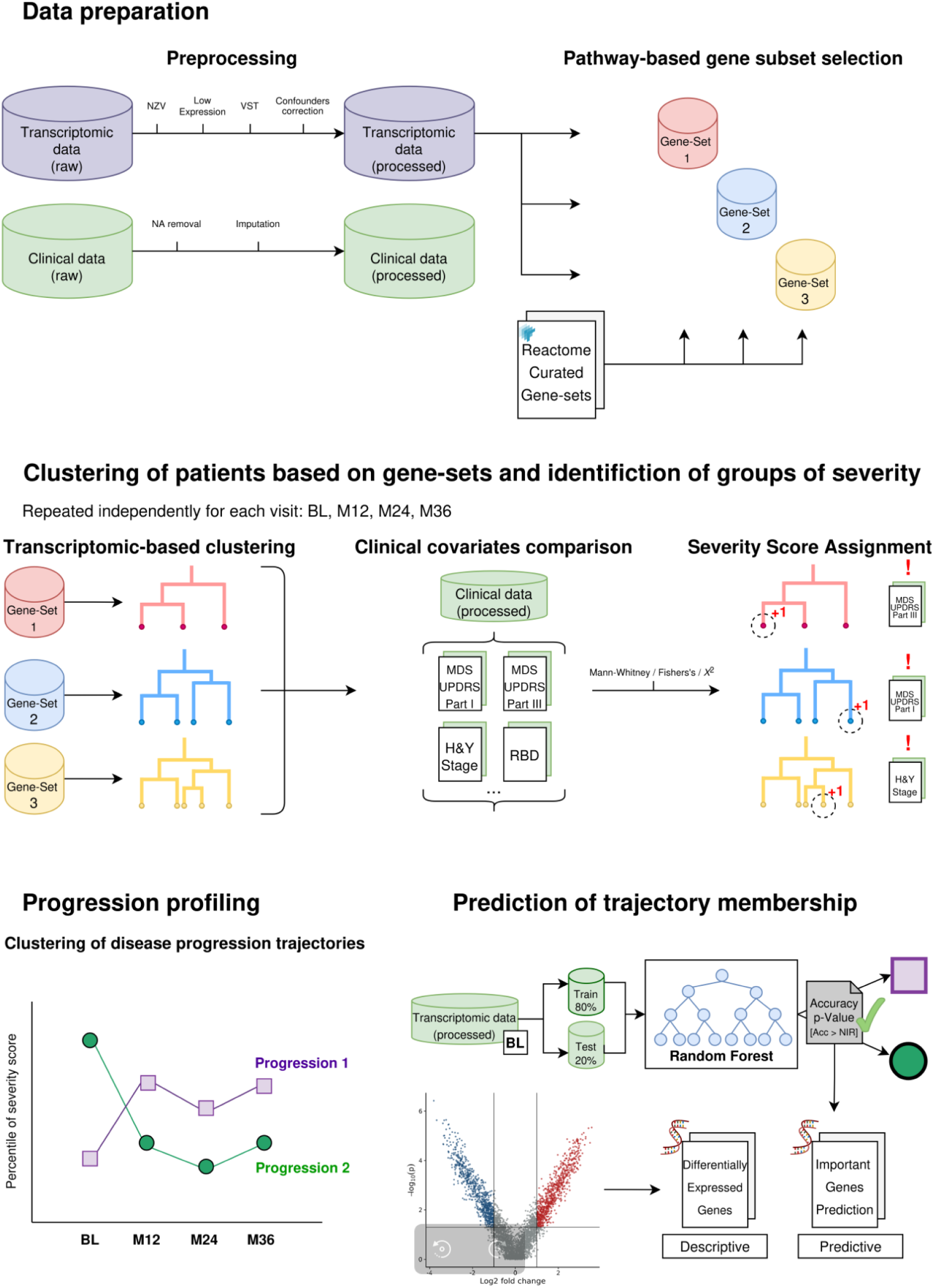
**Schematic Representation of the Study Workflow**

Expression data were normalized and variance-stabilized using the variance stabilizing transformation (vst) implemented in DESeq2 (v1.42.1)^11^. To identify and correct for technical and biological confounders, principal component analysis and variancePartition (v1.32.5)^12^ were used to identify and quantify the proportion of variance explained by candidate covariates. Covariates were selected for correction based on predefined quantitative criteria (upper quartile ≥ 3%, maximum variance contribution ≥ 75%, or weighted Spearman’s correlation ≥ 0.025) and adjusted using linear modeling with variancePartition.

### Clinical Data Preprocessing

Clinical data underwent rigorous quality control prior to analysis. Covariates with more than 5% missing values across participants were removed to minimize bias due to excessive imputation. Those with ≤ 5% missingness were imputed using the cohort-wide median, selected for robustness to skewed distributions and outliers.

For baseline assessments, missing values were imputed using data from the screening visit when available, consistent with the PPMI protocol specifying that baseline and screening visits occur within 60 days^4^. Follow-up visits were not imputed across time points to preserve temporal independence. Following imputation, datasets were reviewed for internal consistency. Static variables exhibiting temporal variability were corrected, and extreme values (>3 standard deviations from the mean) were manually inspected to distinguish valid clinical observations from data entry errors.

### Pathway-based gene set selection

To enhance biological interpretability, we adopted a pathway-based gene sets selection strategy using curated pathway databases. Gene sets were retrieved from the Reactome Curated Pathways collection via the msigdbr package (v24.1.0)^13^.

A strategy based on hierarchical clustering of pathway gene overlap was employed to select a non-redundant set of pathways for downstream analysis (Supplementary Methods 1), reducing the initial set of 1733 Reactome pathways to a final collection of 336 non-redundant pathways.

### Pathway-specific transcriptomic clustering

For each of the 336 selected pathways, hierarchical clustering of patients was performed based on pathway-restricted gene expression profiles, considering only protein-coding genes. Clustering was conducted using Ward’s method as implemented in the hclust function in R^14^, with similarity quantified using the absolute Pearson correlation between samples and transformed into a dissimilarity measure 1 − |𝑐𝑜𝑟𝑟𝑒𝑙𝑎𝑡𝑖𝑜𝑛|.

Cluster solutions were generated for k values ranging from 2 to 20. An upper limit of k=12 was determined based on optimal predictive performance validation (Section 4.8).

Pathway gene set sizes ranged from 5 genes (R-HSA-9667769) to 1154 genes (R-HSA-73857).

### Identification of pathway-associated patient groups and severity score generation

In order to stratify patients by disease severity and progression, pathway-specific clusters (section 4.5) were pairwise compared across clinically relevant covariates to identify significant differences in disease severity. Continuous covariates were analyzed using the Mann–Whitney U test, while ordinal and categorical covariates were analyzed using Fisher’s exact test or the *χ*² test, as appropriate. P-values were Bonferroni-corrected for the number of covariates and cluster comparisons.

When significant differences were detected, patients belonging to the cluster with the more severe clinical profile for a given covariate received a +1 score. Clinical covariates were grouped into motor and non-motor domains according to established clinical frameworks^15,16^ (Supp. Table 1). For each patient and visit, severity signals were summed across pathways within each domain, yielding the Unsupervised Transcriptomic Severity Score (UTSS), computed separately for motor and non-motor domains.

### Longitudinal disease progression trajectory analysis

UTSS values across all available visits were assembled into patient-specific severity profiles {𝑆𝑖0, 𝑆 𝑖1, 𝑆 𝑖2, 𝑆 𝑖3}. To ensure comparability across time points, values were transformed into percentile ranks at each visit.

Disease progression trajectories were defined using feature vectors combining baseline UTSS and longitudinal changes in percentile rank between visits {𝑆 𝑖𝑜, (𝑆 𝑖1 − 𝑆 𝑖𝑜), (𝑆 𝑖2 − 𝑆 𝑖1 ), (𝑆 𝑖3 − 𝑆 𝑖2)}. K-means clustering was performed using Euclidean distance (stats v4.3.1^14^). The optimal number of clusters (*k* = 2–10) was determined based on downstream predictive performance (Section 4.8).

### Prediction of progression trajectories from baseline transcriptomics

Using baseline transcriptomic data, separate Random forest models were trained to predict motor and non-motor progression trajectories across a range of cluster solutions (k = 2–10). UTSS pathway genes served as predictors and were filtered for high correlation (Pearson *r* > 0.95) using training data only, with the same filter applied to test data to prevent leakage. Models were implemented using the *ranger* package (v0.17.0)^17^.

Data were split into training and test sets (80/20), preserving trajectory distribution. Hyperparameter tuning was performed via 5-fold cross-validation, optimizing mtry (10, 30, 60) with 500 trees, splitrule to “gini”, and min.node.size to 10. Model performance was assessed on independent test sets using overall accuracy, with significance tested against the No Information Rate.

### Identification of progression-associated transcriptomic features

To detect the most relevant genes in predicting progression trajectories, gene importance was quantified using permutation-based decreases in model accuracy, and genes were ranked according to their mean importance. The optimal number of predictive genes was selected using a curvature-based elbow method applied to the ranked importance distribution (Supp. Fig. 3).

To complement the non-linear feature selection provided by random forest, differential expression analysis was performed at baseline using DESeq2 (v1.42.1)^11^. For each progression trajectory, gene expression was compared against all other trajectories. Genes were considered significant if they met an adjusted false discovery rate (FDR) < 0.05 (Benjamini–Hochberg correction) and an absolute log₂ fold change > 0.5.

### Functional enrichment analysis of Progression-Associated Transcriptomic Features

Progression-associated gene sets for motor and non-motor domains were functionally characterized using over-representation analysis. Enrichment testing was performed using the compareCluster function (v4.14.3)^18^ on Gene Ontology Biological Process. Enriched terms were considered statistically significant at FDR < 0.05 following Benjamini–Hochberg correction.

### Model validation across genetic and external cohorts

#### Genetic and healthy control cohorts

To evaluate generalizability, linear regression models were trained in the sPD cohort using clinical covariates contributing to the UTSS as predictors and UTSS values as outcomes. Separate models were fitted for motor and non-motor domains at each visit using the lm function in R^14^. These models were then applied to genetic PD cohorts (*GBA*, *LRRK2*, *SNCA*) and HC. Differences in UTSS distributions between groups and the sPD reference cohort were evaluated using the Mann–Whitney U test.

#### PDBP Sporadic cohort

The full analytical pipeline was independently applied to the sporadic PD cohort from the PDBP study. Identical preprocessing, normalization, and modeling procedures were used, with analyses adapted to the PDBP visit schedule.

## Results

### Baseline transcriptomic signatures stratify PD patients by progression trajectory

To characterize longitudinal heterogeneity in Parkinson’s disease progression, we analyzed disease trajectories in 160 sporadic PD patients derived from longitudinal UTSS values, integrating baseline severity and within-cohort percentile changes across visits (Section 3.8).

Two progression trajectories were identified in the non-motor (Figure 2A) and motor (Figure 2B) domains. In the non-motor domain, Cluster 1 (87 patients) showed a more stable course, while Cluster 2 (73 patients) had more pronounced percentile fluctuations. In the motor domain, Cluster 1 (76 patients) exhibited higher baseline severity and greater M0–M12 instability, whereas Cluster 2 (84 patients) had more stable progression.

**Figure 2.**
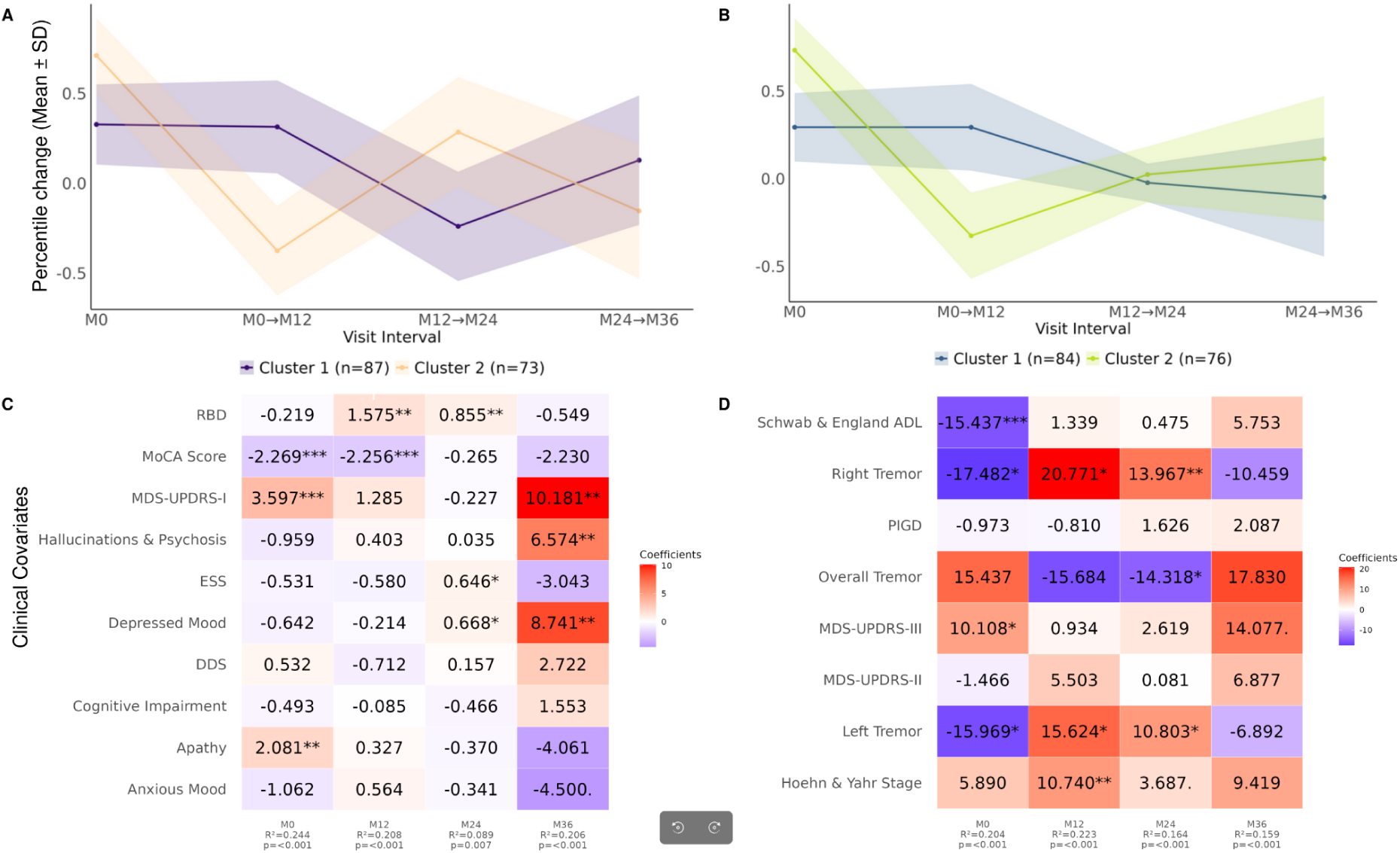
Motor and non-motor progression trajectories and their independent clinical determinants derived from longitudinal transcriptomic severity scores. (A–B) Trajectories were identified using longitudinal severity percentiles. (A) In the non-motor domain, two distinct clusters emerged: Cluster 1 (n = 87) exhibited abrupt percentile changes across visits, whereas Cluster 2 (n = 73) followed a more stable course with less pronounced variation. (B) In the motor domain, Cluster 1 (n = 76) showed higher baseline severity and unstable progression between M0 and M12, while Cluster 2 (n = 84) displayed more stable trajectories across all visits. Baseline transcriptomic profiles were subsequently used to predict trajectory membership, achieving high accuracy for motor progression and significant accuracy for non-motor progression, underscoring the predictive value of molecular signatures for future clinical heterogeneity. (C–D) Independent clinical determinants of transcriptomic severity scores were identified using multivariate linear models. (C) In the non-motor domain, predictors shifted over time: at M0, MDS-UPDRS-I and Apathy were strong positive predictors, whereas MoCA score was negatively associated. At M12, RBD and MoCA remained dominant contributors. By M24, effect sizes decreased overall, although RBD persisted alongside smaller contributions from ESS and Depressed mood. At M36, MDS-UPDRS-I, Depressed mood, and Hallucinations & Psychosis emerged as the principal positive predictors, while Anxious mood showed a negative coefficient. (D) In the motor domain, baseline severity was positively driven by MDS-UPDRS-III, with Right and Left tremor and Schwab & England ADL showing inverse effects. At M12, tremor subcomponents predominated, with Right tremor exerting the strongest positive effect. At M24, lateralized tremors remained positively associated, whereas Overall tremor showed a negative coefficient. By M36, MDS-UPDRS-III became the main positive determinant. For each visit, adjusted R² and overall model p-values are displayed beneath the corresponding visit label, summarizing model fit. Significance levels of individual predictors are indicated as *** p < 0.001; ** p < 0.01; * p < 0.05; . p < 0.1.

**Figure 3.**
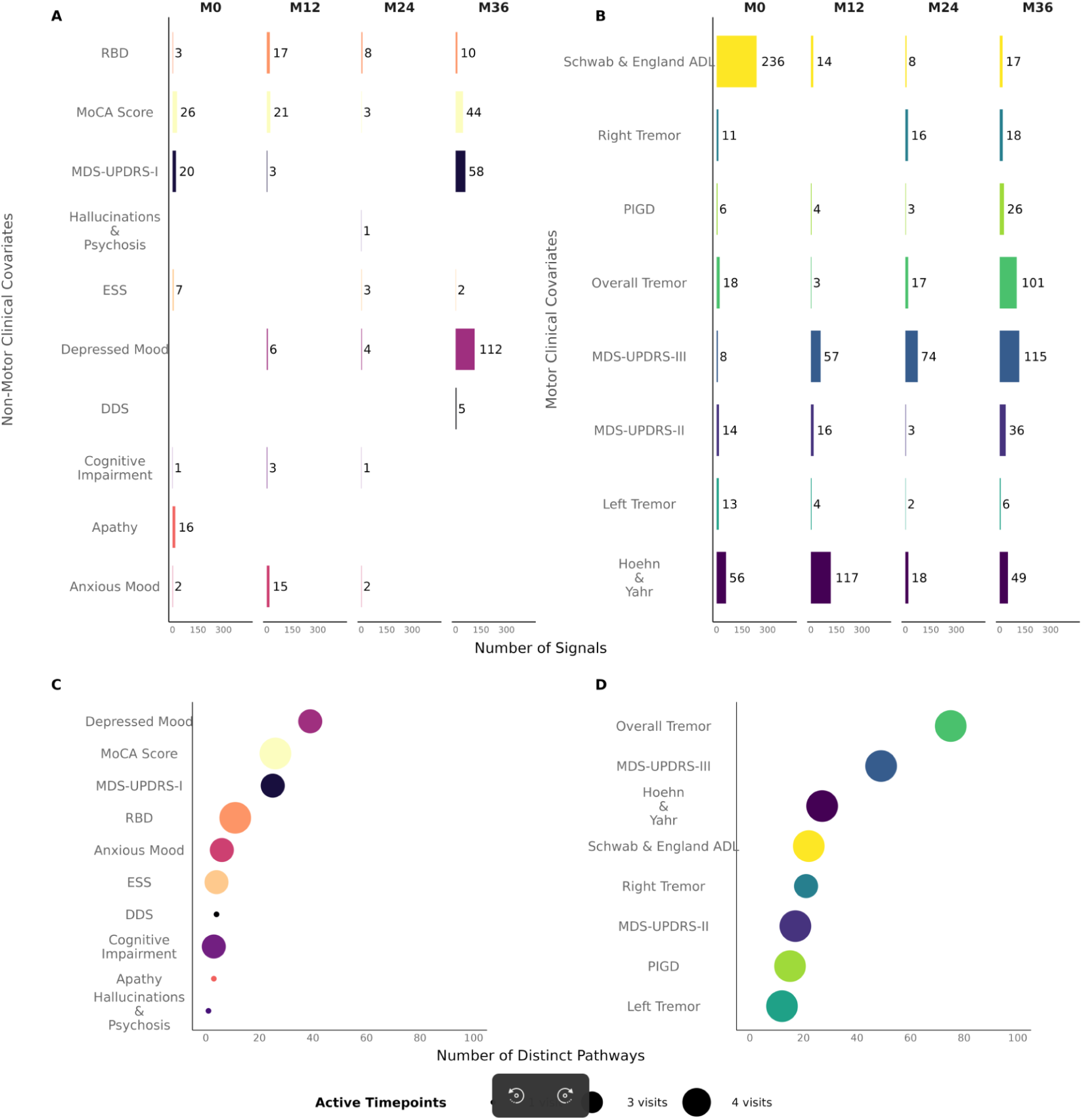
Pathway-level correlates of clinical heterogeneity captured by the transcriptomic severity score over time. (A) Non-motor domain: The number and composition of pathway-clinical covariate associations varied across visits, with “MDS-UPDRS-I”, “MoCA score”, and “Apathy” dominating at baseline, “RBD” and “MoCA score” at M12, and “Depressed mood” emerging as the primary feature by M36. Other features, including hallucinations, cognitive impairment, and ESS, contributed consistently fewer signals. (B) Motor domain: At baseline, “Schwab & England ADL” dominated associations, with “Hoehn & Yahr” and other motor features with lesser contributions; over time, “Hoehn & Yahr” and “MDS-UPDRS-III” gained prominence, and by M36, “Overall tremor” and “MDS-UPDRS-III” were the most represented features, while “Schwab & England ADL” and “MDS-UPDRS-II” remained detectable. (C) Overall non-motor pathway persistence: Fewer unique pathways were associated per feature, with “Depressed mood” showing the highest diversity; in the motor domain. (D) Overall motor pathway persistence: broader representation was observed, with “Overall tremor”, “MDS-UPDRS-III”, and “Hoehn & Yahr” linked to the largest number of distinct pathways.

Random Forest classifiers trained on baseline transcriptomic features achieved robust discrimination of trajectory membership (Supp.Table 3). For motor progression, the optimal two-cluster model achieved an accuracy of 0.871 (95% CI: 0.70–0.96), significantly exceeding the No Information Rate (p = 3.5 × 10⁻⁵), with high specificity (0.93), and sensitivity (0.81).

In the non-motor domain, random forest model also demonstrated significant predictive performance (accuracy = 0.77; 95% CI: 0.59–0.90; p = 0.008), with balanced sensitivity (0.7059) and specificity (0.86). Models configured with higher numbers of clusters (k = 3–4) yielded lower or non-significant predictive performance in both domains.

### Pathway-level transcriptomic correlates of clinical features

We next examined how clinical features contributed to UTSS values through pathway-specific transcriptomic clustering (Section 3.6). The distribution of these contributions across covariates and visits is summarized in Figure 3A–B.

In the non-motor domain (Figure 3A), the number and identity of contributing clinical features varied substantially over time. At baseline, MDS-UPDRS-I accounted for the largest number of associations, followed by apathy and cognitive performance (MoCA). At M12, REM sleep behavior disorder (RBD) and MoCA emerged as the dominant contributors. Signal counts declined at M24, whereas at M36 depressed mood became the most prominent contributor, followed by MDS-UPDRS-I and MoCA. Other non-motor features showed comparatively fewer pathway-level associations across visits.

In the motor domain (Figure 3B), Schwab & England Activities of Daily Living (S&E ADL) dominated baseline associations. At later visits, contributions shifted toward core motor severity measures, particularly Hoehn & Yahr stage and MDS-UPDRS-III, as well as tremor-related features. By M36, MDS-UPDRS-III and tremor measures accounted for the largest proportion of pathway-level associations, while S&E ADL retained a smaller but persistent contribution.

Figures 3C–D summarize the diversity of pathways associated with each clinical feature across visits. Overall, pathway diversity was greater in the motor domain. In the non-motor domain, depressed mood, MoCA, and MDS-UPDRS-I showed the most associations, while most other features linked to fewer, visit-specific pathways. In contrast, motor features, particularly tremor, MDS-UPDRS-III, and Hoehn & Yahr stage—were associated with a broader repertoire of pathways.

Pathway contributions to UTSS patterns evolved over time. At baseline, transport and metabolic processes dominated, including SLC-mediated transmembrane transport, vitamin B12 metabolism, lipid biosynthesis, tRNA aminoacylation, and hemostasis. At intermediate visits, enrichments shifted toward RNA and protein homeostasis and cellular stress responses, highlighting mRNA decay, protein folding, mitochondrial tRNA modification, gluconeogenesis, and calcium transport. By M36, signaling and immune-related pathways predominated, including EPH–ephrin signaling, CD28 co-stimulation, neutrophil degranulation, and STAT3-and NOTCH-mediated cascades. In contrast, core neuronal and bioenergetic pathways—such as dopamine clearance, choline catabolism, membrane potential regulation, and mitochondrial function—remained consistently enriched, suggesting a persistent contribution to patient stratification. (Supp. Table 2).

### UTSS captures distinct clinical determinants in motor and non-motor domains

To assess the clinical determinants underlying UTSS values, we fitted multivariate linear models at each visit including all relevant clinical covariates as predictors (Figure 4A–B).

**Figure 4.**
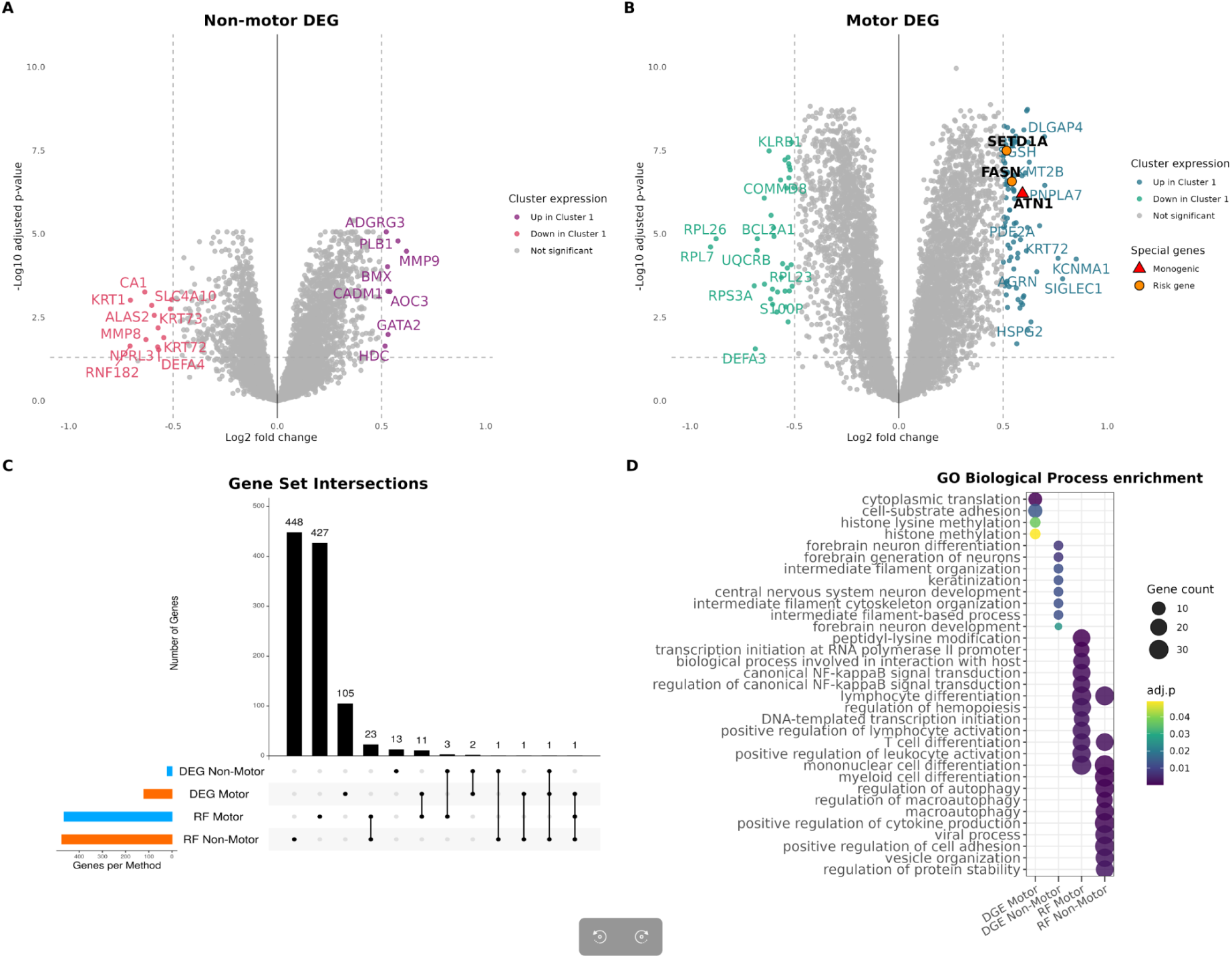
Baseline transcriptomic signatures associated with motor and non-motor progression trajectories. (A) Non-motor domain: Volcano plot showing differential gene expression at baseline across non-motor progression trajectories, revealing a limited transcriptional separation with a small number of significant DEGs (FDR < 0.05, |log₂FC| > 0.5). Genes are colored according to the trajectory in which they are regulated. (B) Motor domain: Volcano plot of baseline DEGs between stable and unstable motor progression trajectories, showing a markedly stronger transcriptomic signal and a larger number of differentially expressed genes. In panels A and B, genes previously associated with Parkinson’s disease, including monogenic and genetic risk genes, are highlighted and labeled. (C) UpSet plot summarizing overlaps between genes identified by DGE and Random Forest (RF) feature selection in both domains. Only minimal overlap was observed between linear (DGE) and non-linear (RF) approaches, highlighting complementary transcriptomic signals associated with progression. (D) GO Biological Process enrichment of DGE- and RF-derived gene sets. Enrichment patterns were strongly method-dependent. DGE-derived signatures were dominated by translation-, adhesion-, chromatin-, and neurodevelopment-related processes, whereas RF-derived signatures in both domains were predominantly immune and inflammatory. Notably, adaptive immune and NF-κB–related processes characterized RF Motor genes, while RF Non-Motor genes showed enrichment for myeloid, cytokine, and stress-response pathways.

In the non-motor domain (Figure 4A), model performance varied across visits, with adjusted R² values of 0.244 at M0, 0.208 at M12, 0.089 at M24, and 0.206 at M36 (all p ≤ 0.05). At baseline, higher UTSS non-motor values were independently associated with greater MDS-UPDRS-I scores, apathy, and lower cognitive performance. At M12, RBD and MoCA score were the primary predictors. By M24, the model’s explanatory power was lowest, with RBD being the sole independent predictor alongside small contributions from ESS and depressed mood. By M36, affective and neuropsychiatric features— depressed mood, hallucinations and psychosis, and MDS-UPDRS-I—dominated the model.

Motor domain models (Figure 4B) were statistically significant at all visits (p ≤ 0.001) and showed relatively stable explanatory power (adjusted R² = 0.16–0.22). At baseline, UTSS motor was positively associated with MDS-UPDRS-III and inversely with tremor laterality and S&E ADL. At subsequent visits, tremor-related measures and Hoehn & Yahr stage were the main predictors. By M36, explanatory power declined, with only marginal associations remaining.

### Progression-associated transcriptomic signatures reveal distinct functional pathways

We next characterized baseline transcriptomic differences between progression trajectories using complementary differential gene expression (DGE) and random forest-based variable importance approaches.

In the non-motor domain, DGE identified 20 differentiating genes, indicating subtle transcriptional separation, while random forest modeling identified 475 predictive genes, with minimal overlap between methods (Figure 5A,C). In the motor domain, differences were more pronounced: DGE identified 121 and random forest selected 465 genes, again with limited overlap (Figure 5B–C).

**Figure 5.**
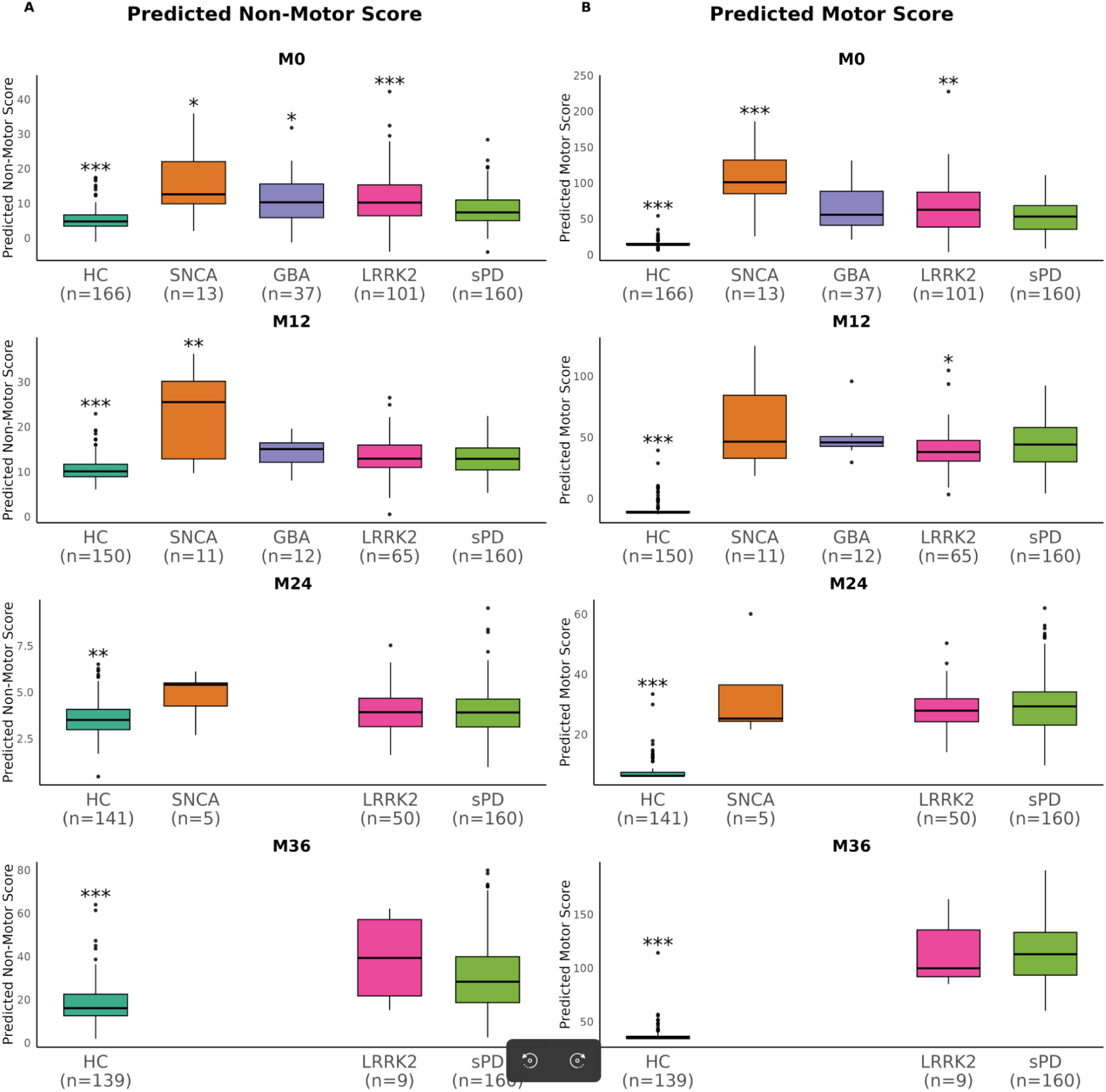
Generalization of clinical severity prediction models to independent PPMI cohorts. (A) Non-motor domain: Predicted non-motor severity scores displayed greater dispersion and overlap between cohorts. Although HC generally showed lower severity than PD groups, substantial overlap was observed, particularly among sPD, LRRK2, and GBA. The SNCA cohort tended to exhibit higher median non-motor severity, though variability increased at later visits, reflecting reduced discriminatory power relative to the motor domain. (B) Motor domain: Boxplots show predicted transcriptomic severity scores across sporadic PD, genetic PD cohorts (SNCA, LRRK2, GBA), and healthy controls (HC) at each visit. Predicted motor severity was consistently higher in PD-related cohorts than in HC across all time points, with the SNCA cohort showing the highest median severity, followed by GBA and LRRK2, and sPD exhibiting intermediate values. These patterns were stable over time, indicating robust generalization of the motor models.

Functional enrichment revealed domain-specific signatures (Figure 5D). In the motor domain, DGE genes were enriched for cytoplasmic translation, cell–substrate adhesion, and histone methylation, whereas random forest genes highlighted regulatory and immune mechanisms, including transcriptional initiation, NF-κB signaling, and lymphocyte differentiation.

In the non-motor domain, DGE-based enrichment was primarily associated with neurodevelopmental and structural processes (e.g., forebrain neuron differentiation, CNS development), whereas RF-selected genes showed strong enrichment for immune and stress-response pathways, including mononuclear and myeloid differentiation, cytokine production, autophagy, vesicle organization, and protein stability.

Several identified genes overlapped with prior genetic evidence to Parkinson’s disease or clinically related movement disorders. In the motor domain, these included genes causing monogenic movement disorders with overlapping clinical features (*ATN1*, *HTT*, *PLA2G6*), as well as GWAS susceptibility genes^19,20^ (*FGD4*, *SPTSSB*, *SCARB2*, *SETD1A*, *FCGR2A*, *FASN*, *ASH1L*, *CCT3*). In the non-motor domain, overlap was more prominent, encompassing canonical monogenic drivers (*SNCA*, *SYNJ1*, *RAB39B*, *PDGFB*) and GWAS-implicated genes (*BAG3*, *IP6K2*, *CTSB*, *SEMA4A*, *INPP5F*, *AREL1*, *FCGR2A*).

### Clinical models generalize to genetic PD and healthy control cohorts

To assess generalizability, UTSS predictive models trained in sporadic PD were applied to genetic PD cohorts (*SNCA*, *LRRK2*, *GBA*) and healthy controls (HC) (Figure 6).

In the non-motor domain, HC consistently exhibited lower predicted UTSS values compared with PD cohorts at all visits (p < 0.05). Substantial overlap was observed among sporadic and genetic PD groups, although *SNCA* carriers tended to show higher median severity, particularly at earlier visits. In the motor domain, predicted UTSS values were consistently higher in PD cohorts than in HC across all visits, with minimal overlap. HC differed significantly from sporadic PD at all visits, and *SNCA* carriers showed higher motor severity at baseline.

### Independent replication in the PDBP cohort validates the UTSS framework

To assess replication, we applied the UTSS pipeline to the independent PDBP cohort, an external longitudinal study with harmonized blood transcriptomic and clinical measures. Consistent with PPMI, this analysis identified two non-motor (n=119, 126) and two motor (n=125, 120) progression trajectories (Supp. Fig. 4).

Clinical contributions to UTSS (Supp. Fig. 5) differed from PPMI, with early contributions from hallucinations and psychosis, depressed mood, and cognitive impairment. UTSS-M was dominated by tremor measures and PIGD throughout follow-up, while the contribution of MDS-UPDRS-III decreased over time.

UTSS-generated pathways showed high overlap between PPMI and PDBP (Szymkiewicz–Simpson index = 0.85; Supp. Table 4), with shared terms reflecting canonical neurodegenerative processes, including dopamine metabolism, mitochondrial dysfunction, autophagy, immune signaling, and metabolic dysregulation.

Transcriptomic analyses in PDBP identified progression-associated genes previously implicated in monogenic parkinsonism or related neurodegenerative disorders (e.g., *PANK2*, *SNCA*, *DNAJC6*), as well as GWAS risk loci (e.g., *GPNMB*, *WNT3*, *CTSB*). Several signals overlapped with PPMI (e.g., *SNCA*, *CTSB*, *FCGR2A*), supporting cross-cohort consistency.

At the pathway level, non-motor progression in PDBP was enriched for immune-related processes, including leukocyte activation and cytokine signaling, consistent with PPMI. In contrast, motor progression in PDBP was characterized by enrichment in ion transport, sensory perception, and hypoxia-related pathways, differing from the vesicle and transcriptional signatures predominating in PPMI. (Supp. Fig 9).

## Discussion

Using longitudinal data from the Parkinson’s Progression Markers Initiative, we show that blood-based transcriptomic pathways are associated with distinct motor and non-motor progression trajectories, capturing clinically meaningful heterogeneity in sporadic PD from early stages.

Our analysis reveals that baseline blood transcriptomic profiles delineate distinct motor and non-motor progression trajectories in sporadic PD from disease onset. This early divergence suggests inherent biological heterogeneity at initial stages, which diminishes over time as motor phenotypes converge—a pattern consistent with the phenotypic convergence previously reported.^21,22^

We found that motor progression trajectories were supported by pathway-level signals aligned with established PD mechanisms, including sustained dysregulation of dopaminergic and cholinergic signaling and persistent mitochondrial function pathways. While these processes are classically studied in the central nervous system, systemic metabolic and immune alterations may allow components of these pathways to be captured in peripheral blood transcriptomics. These findings are consistent with evidence implicating neurotransmitter imbalance and mitochondrial dysfunction in distinct motor phenotypes^23–25^.

We also show an association between cobalamin transport and metabolism and Hoehn & Yahr stage progression, consistent with evidence linking vitamin B12 deficiency to a more progressive PD phenotype^26^. These pathway-level associations provide biological support for the observed motor progression trajectories.

In contrast, we observed that non-motor progression was supported by a distinct set of pathway-level transcriptomic signals. Particularly, trajectories with greater non-motor burden and cognitive decline showed the strongest association with Chondroitin sulfate/dermatan sulfate metabolism, which have been linked to neuroinflammatory responses and oxidative stress^27^. Immune dysregulation was further suggested by enrichment of the Nef-mediated CD8 downregulation pathway, indicating potential involvement of peripheral immune processes in cognitive decline^28^. Additional persistent signals related to ionic homeostasis and PKA-mediated phosphorylation were also observed, consistent with prior literature linking these processes to neuroprotective mechanisms ^29–31^.

We extend prior genetic studies by showing that the implicated pathways—primarily involving immune processes, metabolic functions, and neurodevelopmental signaling—partially overlap with pathways enriched for genes identified in PD genome-wide association studies and monogenic parkinsonism ^32^. This convergence was observed at the pathways-enrichment level rather than at single-gene resolution and was replicated in the independent PDBP cohort, supporting the robustness of these associations.

Our results show that motor and non-motor progression in PD are driven by largely independent transcriptomic programs, reflecting their clinical dissociation. Motor trajectories involved stronger transcriptional changes in pathways like cytoplasmic translation and vesicle-mediated transport, while non-motor trajectories showed subtler, immune-related alterations. Both domains were associated with PD-relevant genes (i.e. S*NCA*, *BAG3*), including monogenic drivers and susceptibility loci. Integrating DGE with Random Forest models revealed progression-relevant signals beyond population-level differences, underscoring the need to assess motor and non-motor progression separately.

We further showed that the proposed framework generalizes across clinically and genetically distinct PD populations. When applied to genetic cohorts within PPMI, the predicted progression profiles closely reflected known phenotypic differences. Specifically, motor and non-motor trajectories predictions for *LRRK2* mutation carriers aligned with those observed for sPD, consistent with their overlapping clinical presentation. *SNCA* carriers were predicted to have significantly higher non-motor burden, aligning with their established phenotype of greater cognitive and psychiatric involvement.^33,34^ Replication in the independent PDBP cohort confirmed:

i. the presence of two motor and two non-motor progression trajectories, (ii) similar longitudinal patterns of convergence in motor trajectories and relative stability in non-motor trajectories, and (iii) enrichment of overlapping pathways-level signals, including dopamine clearance pathways consistently associated with Hoehn & Yahr stage progression.

Several limitations of this study warrant consideration. First, our findings were replicated in genetic cohorts, the sample sizes, particularly for genetic subgroups (e.g., *SNCA*), remain modest, which may affect the precision of subtype-specific predictions. Second, the potential confounding effect of the medication exposure, present in baseline PDBP but not in PPMI, may influence transcriptomic profiles and progression patterns. Finally, the blood transcriptome serves as a systemic proxy, and the precise relationship between peripheral signals and central nervous system pathology requires further mechanistic validation. The proposed framework is not intended to replace clinical judgment but rather to complement existing clinical assessments by providing additional prognostic information.

In conclusion, our study provides evidence that blood-based transcriptomic profiling can capture clinically meaningful heterogeneity in sporadic Parkinson’s disease by identifying distinct motor and non-motor progression trajectories from early disease stage. These trajectories reflect differential longitudinal courses that are not readily apparent at baseline and are supported by biologically reported pathway-level signals. By enabling early, domain-specific prognostic stratification, our framework provides a clinically interpretable approach to understanding disease progression in PD. While further validation is required, specifically in prospective and interventional trial cohorts, this work represents an important step forward improving patient stratification and potentially optimizing the design of clinical trials targeting disease progression.

## Data sharing

The data that supports the findings of this study are available in the supplementary material of this article.

## Author’s Roles

**(1) Research Project**: A. Conception, B. Organization, C. Execution; **(2) Statistical Analysis**: A. Design, B. Execution, C. Review and Critique; **(3) Manuscript**: A. Writing of the First Draft, B. Review and Critique. All authors approved the final version of the manuscript.

J.Ñ.: 1A, 1C, 2B, 3A, 3B. A.G.: 2A, 2C.

G.R.: 1C.

H.M.: 1B.

M.R.: 1B, 3B.

J.T.P.: 1A, 1B, 1C.

J.A.B.: 1A, 1B, 1C, 2C, 3B.

A.-L.G.-M.: 1C, 3B.

## Code availability

Code repository. https://github.com/JaimeBioR/pathway-pd-progression

## Data Availability

All data produced in the present study are available upon reasonable request to the authors

